# Effects of boosted mRNA and adenoviral-vectored vaccines on immune responses to omicron BA.1 and BA.2 following the heterologous CoronaVac/AZD1222 vaccination

**DOI:** 10.1101/2022.04.25.22274294

**Authors:** Nungruthai Suntronwong, Sitthichai Kanokudom, Chompoonut Auphimai, Suvichada Assawakosri, Thanunrat Thongmee, Preeyaporn Vichaiwattana, Thaneeya Duangchinda, Warangkana Chantima, Pattarakul Pakchotanon, Jira Chansaenroj, Jiratchaya Puenpa, Pornjarim Nilyanimit, Donchida Srimuan, Thaksaporn Thatsanatorn, Natthinee Sudhinaraset, Nasamon Wanlapakorn, Yong Poovorawan

## Abstract

**Background:** The coronavirus 2019 omicron variant has surged rapidly and raises concerns about immune evasion because it harbors mutations even in individuals with complete vaccination. Here, we examine the capability of the booster vaccination to induce neutralizing antibodies (NAbs) against omicron (BA.1 and BA.2) and T-cell responses.

**Methods:** A total of 167 participants primed with heterologous CoronaVac/AZD1222 were enrolled to receive AZD1222, BNT162b2, or mRNA-1273 as a booster dose. Reactogenicity was recorded. Binding antibody, neutralizing antibody (NAb) titers against omicron BA.1 and BA.2, and total interferon gamma (IFN-γ) post-booster responses were measured.

**Results:** A substantial loss in neutralizing potency to omicron variant was found at 4 to 5 months after receiving the heterologous CoronaVac/AZD1222. Following booster vaccination, a significant increase in binding antibodies and neutralizing activities toward delta and omicron variants was observed. Neutralization to omicron BA.1 and BA.2 were comparable, showing the highest titers after boosted mRNA-1273 followed by BNT162b2 and AZD1222. Notably, boosted individuals with mRNA vaccines could induce T cell response. Reactogenicity was mild to moderate without serious adverse events.

**Conclusions:** Our findings highlight that the booster vaccination could overcome immunity wanes and provide adequate NAbs coverage against omicron BA.1 and BA.2.

## INTRODUCTION

As of November 2021, the severe acute respiratory syndrome coronavirus 2 (SARS-CoV-2) omicron (B.1.1.529) variant quickly surged worldwide and raised concern about immune evasion [1]. The omicron variant is characterized by many mutations in the spike protein. Among these, fifteen mutations located on the receptor-binding domain (RBD), which are responsible for interactions with the angiotensin-converting enzyme 2 (ACE2) receptor, and eight amino acid changes are found at the N-terminal domain (NTD) [2]. In a study of mutated RBD profiles, various omicron mutations capable of escaping human neutralizing antibodies (NAbs) were found to contain epitopes overlapping the ACE2 binding motif, indicating evasion of immunity and reduction in vaccine effectiveness [3]. Furthermore, mutations in NTD related to partial escape from the Nabs have been reported [4]. These findings are consistent with other clinical data demonstrating that the emergence of the omicron variant has led to an increase in the risk of reinfection [5]. During the ongoing viral evolution, the omicron variant has been divided into four subvariants, including BA.1, BA.1.1, BA.2, and BA.3 [6]. Among them, BA.1 surged earlier and became the dominant subvariant circulating worldwide. However, the BA.2 subvariant has recently increased in multiple countries and appears to be more transmissible than BA.1 [7]. Previous studies suggest that BA.1 and BA.2 are highly resistant to neutralization by monoclonal antibody therapy and vaccine-induced immunity [8,9].

Another concern is the waning immunity that occurs over time. A previous study indicates that the IgG antibodies declined a consistent rate at six months after second dose of the mRNA vaccination while neutralizing antibodies declined rapidly over the first three months followed by a relatively slower decrease after that point [10]. A reduction in NAbs is related to an increased risk of symptomatic SARS-CoV-2 infection and reduced vaccine effectiveness [11].

Moreover, omicron variants were poorly or not at all neutralized in the sera sampled five months after completing the two-dose BNT162b2 or AZD1222 vaccination courses [12]. Due to the emergence of omicron and waning immunity, a booster vaccination program has been implemented in many countries [13,14]. Thus, data on boosting immunity against omicron variant are needed.

Besides humoral immunity, cell-mediated immunity also plays an essential role in limiting the SARS-CoV-2 infection [15] and reducing disease severity in acute coronavirus 2019 (COVID-19) patients [16]. The SARS-CoV-2-specific T-cells persist at least six months after receiving the two-dose regimen of either mRNA or adenoviral-vectored vaccines, whereas the levels of NAb severely declined [17]. Furthermore, SARS-CoV-2 specific T-cells induced by vaccination or previous infection are highly cross-reactive with the omicron variant, and the omicron-cross reactive T cells exhibiting the polyfunctional profiles were not significantly different compared to ancestral strain and other SARS-CoV-2 variants [18]. These findings indicate that a few mutations in the spike can minimally affected T-cell recognition.

Seven vaccines have now been authorized in Thailand: (1) inactivated CoronaVac, (2) BBIBP-CorV, (3) adenoviral-vectored ChAdOx1-S/AZD1222, (4) Ad26.COV2.S, (5) mRNA-based BNT162b2, (6) mRNA-1273, and (7) protein-based NVX-CoV2373 vaccines [19]. However, the vaccine effectiveness and capability of inducing immune responses differ with different types of vaccines and are affected by emerging variants [20]. Our previous study indicated that mRNA vaccine-or AZD1222-boosted individuals after a two-dose CoronaVac course elicited a higher immune response than in those receiving boosted inactivated vaccines [21]. In the COV-BOOST trial, the heterologous boost after either two-dose of AZD1222 or BNT162b2 prime showed an increased humoral and cell-mediated immune response compared to homologous booster vaccination, although the reactogenicity was increasing in some heterologous boosted combination [22]. However, information about the effects of booster vaccinations on safety and immune responses against omicron variants following heterologous primed has been limited.

Due to the limited vaccine supply, Thailand has administered the heterologous CoronaVac followed by AZD1222 vaccination as an alternative regimen for combatting the delta variant. This regimen could induce a higher immune response than the homologous CoronaVac regimen [23]. However, the antibodies wane over time, and the emergence of the omicron variant has raised concerns about booster vaccinations. In this study, the safety and capability of inducing NAbs against the omicron variant (BA.1 and BA.2 subvariants) and T-cell responses after receiving AZD1222, BNT162b2, or mRNA-1273 as a booster dose in individuals who were previously vaccinated with the heterologous CoronaVac/AZD1222 regimen were evaluated. These findings support the policymakers’ choices about which booster vaccines to use in the population to overcome waning immunity and prevent breakthrough infections during the recent emergence of the SAR-CoV-2 variants.

## MATERIALS AND METHODS

### Study design

In a cohort study, 167 individuals who had received CoronaVac/AZD1222 vaccination at least 4 to 5 months earlier and with no history of SARS-CoV-2 infection were enrolled. The participants were offered immunization with one of three approved vaccines, including AZD1222, BNT162b2, or mRNA-1273 vaccines. The cohort study started between November 30, 2021 and January 24, 2022. The participants received adverse events (AEs) documents to record reactogenicity within seven days after receiving their booster dose. Blood samples were collected before vaccination (day 0) and on days 14 and 28 after booster vaccination. The study protocol was reviewed and approved by the Institutional Review Board of the Faculty of Medicine, Chulalongkorn University (IRB numbers 871/64) and performed under the Declaration of Helsinki and Good Clinical Practice principles. This study was registered with the Thai Clinical Trials Registry (TCTR20211120002). All participants signed a written consent before being enrolled in this study.

### Study vaccines

The study vaccines used as booster doses included AZD1222 (AstraZeneca, Oxford, UK) [24], BNT162b2 (Pfizer-BioNTech Inc., New York City, NY) [25], and mRNA-1273 (Moderna Inc., Cambridge, MA) [26]. All vaccines were designed using the SARS-CoV-2 spike of ancestral strain as a template.

### Measurement binding antibody and neutralizing activity using ELISA-based assay

All sera samples measured anti-RBD IgG and anti-nucleocapsid (N) IgG using enzyme-linked immunosorbent assays (Abbott Diagnostics, Abbott Park, IL). The anti-RBD IgG was expressed as a binding antibody unit (BAU/mL), and a value ≥ 7.1 was considered positive. Anti-N IgG reported as S/C ratio (optical density [OD] sample divided by calibrator) with a value ≥ 1.4 was considered positive. In addition, a subset of samples was randomly selected to perform surrogate virus neutralization assay (sVNT) against delta and omicron variants using a cPass™ SAR-CoV-2 neutralizing antibody detection kit (GenScript Biotech, Piscataway, NJ) as previously described [23]. Neutralizing activity was reported as the percentage of inhibition (inhibition [%] = [1 − OD value of sample/average OD of negative control] × 100). A value ≥ 30% was defined as positive, indicating the presence of neutralizing antibodies. The lower limit of detection was set as 0% inhibition.

### Focus Reduction Neutralization test (FRNT50)

Live SARS-CoV-2 neutralizing antibody titers in a subset of serum samples were determined using a 50% focus reduction neutralization test (FRNT50) against omicron BA.1 (accession number: EPI_ISL_8547017) and BA.2 (EPI_ISL_11698090) subvariants. Briefly, heat-inactivated sera were used to prepare serial dilutions starting from 1:10 to 1:7,290 and incubated with live virus for 1 h at 37 °C. The virus–sera mixtures were transferred to monolayers of Vero cells in a 96-well plate and incubated for 2 h. Foci development evaluation and infected cell counting were performed as previously described [27]. The focus reduction percentage for an individual sample was calculated, and the half-maximal inhibitory concentration (IC50) was evaluated using PROBIT software. If no neutralization was observed, the FRNT50 was set as 10, which is one dilution step below the lower limit of detection (dilution 1:20).

### Quantification of interferon-gamma response

The SARS-CoV-2-specific T-cell response was evaluated by using a commercially available IFN-γ release assay in whole blood according to the manufacturer’s instructions (QuantiFERON, Qiagen, Hilden, Germany). Heparinized whole blood was incubated with different antigens, including negative (Nil), positive (Mitogen) and two different SARS-CoV-2 antigens (Ag1 and Ag2). The Ag1 tube was coated with S1subunit (RBD) peptides with CD4+ stimulation, and Ag2 contained S1+S2 peptides for CD4+ and CD8+ stimulation. After stimulation, total IFN-γ production was measured as previously described [23]. The results were calculated from a standard curve and expressed as IU/mL after subtraction of the Nil control. The total IFN-γ with a value ≥ 0.15 IU/mL and ≥ 25% of Nil were considered a positive response against SARS-CoV-2.

### Statistical analyses

Baseline characteristics were expressed as number or percentage and median with interquartile ranges (IQRs). The levels of binding antibody and NAbs were presented as geometric mean titers (GMT). A comparison of log-transformed data was determine using one-way analysis of variance (ANOVA) with Bonferroni adjustment. The Kruskal–Wallis test with Dunn’s post hoc correction and Mann-Whitney test were used for unpaired samples, while the Friedman and Wilcoxon signed-rank tests were used for paired samples in cases in which data were not normally distributed. Spearman’s R was used to determine the correlation. A *P*-value < 0.05 was considered statistically significant. All statistical analyses were performed using SPSS v23.0 (IBM Corp, Armonk, NY). Figures were generated using GraphPad Prism v9.0 (GraphPad, San Diego, CA) and R version 3.6.0 (R Foundation, Vienna, Austria). Details of statistical analysis for each experiment are described in the figure legends.

## RESULTS

### Study participants

A total of 167 vaccinated individuals who received heterologous CoronaVac/AZD1222 were enrolled to receive AZD1222 (n = 60), BNT162b2 (n = 55), and mRNA-1273 (n = 52) with a booster interval ranging from 4 to 5 months post-second dose (Figure 1A). Overall, participants included 83 (49.7%) women and 84 (50.3%) men with ages ranging from 19 to 64 years. The mean age of participants who received AZD1222 was 41.2 years (IQR: 38.5–43.9), BNT162b2 39.0 years (IQR: 36.4–41.6), and mRNA-1273 43.9 years (IQR:40.9–46.8) as booster doses were not significantly different (Table 1). Additionally, no statistically significant differences were observed in the time intervals between the first and the second doses for any group. However, the median interval between the second and third dose in AZD1222 (125 days, IQR: 118–134.5) was slightly shorter (but not statistically significant) than BNT162b2 (130 days, IQR: 110–141) and mRNA-1273 (131 days, IQR:102.3–133).

**Table 1:** Characteristics of participants in the study.

| Characteristics | Total<br>(n=167) | AZD1222<br>(n = 60) | BNT162b2<br>(n = 55) | mRNA-1273<br>(n = 52) | p-value |
| --- | --- | --- | --- | --- | --- |
| Sex (n, %) |  |  |  |  |  |
| female | 83 (49.7%) | 37 (61.7%) | 26 (47.3%) | 20 (38.5%) | $p = 0.045^a$ |
| male | 84 (50.3%) | 23 (38.3%) | 29 (52.7%) | 32 (61.5%) |  |
| Age in years (mean, IQR) | 41.3 (35-48) | 41.2 (38.5-43.9) | 39.0 (36.4-41.6) | 43.9 (40.9-46.8) | $p = 0.052^b$ |
| Interval between 1<sup>st</sup> and 2<sup>nd</sup> dose (median, IQR) | 27 (21-28) | 24 (21-28) | 27 (21-28) | 27 (24.3-28) | $p = 0.332^c$ |
| Interval between 2<sup>nd</sup> and 3<sup>rd</sup> dose (median, IQR) | 130 (110-135) | 125 (118-134.5) | 130 (110-141) | 131 (102.3-133) | $p = 0.281^c$ |

<sup>a</sup> represents the association in sex distributed between groups using Chi-square test. <sup>b</sup> represents the mean different in age between groups using one-way ANOVA. <sup>c</sup> represents the comparison in dosing interval between groups using Kruskal-Wallis H test.

**Figure 1:**
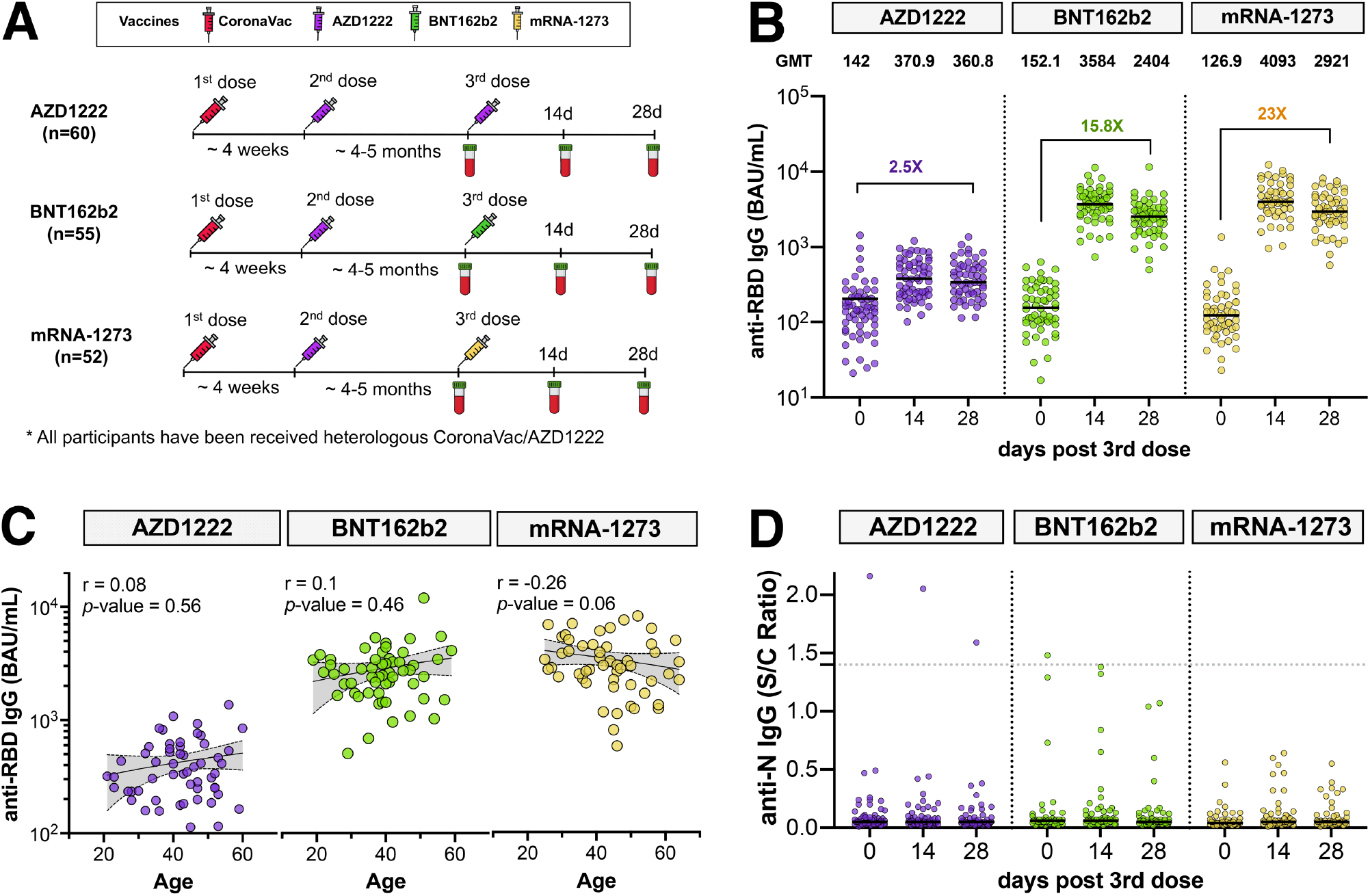
Study design and measurement of severe acute respiratory syndrome coronavirus 2 (SARS-CoV-2)-specific binding antibody responses. Schematic depicting a total of 167 vaccinated individuals with heterologous CoronaVac/AZD1222 enrolled in cohort study. They were assigned to receive booster vaccines, either AZD1222 (*n* = 60), BNT162b2 (*n* = 55), or mRNA-1273 (*n* = 52) and blood samples were collected on days 0, 14, and 28 after booster vaccination **(Panel A**). The anti-receptor binding domain (RBD) IgG (BAU/mL) in sera from boosted individuals with different vaccines, AZD1222 (purple), BNT162b2 (green) and mRNA-1273 (yellow), were compared. **(Panel B)**. Scatter plots shows simple linear fit between anti-RBD IgG and age with Spearman’s R correlation coefficients and *P-*values **(Panel C**). Anti-N IgG was measured at different time points **(Panel D)**. Error bars in **Panels B and D** indicate the geometric mean titers (GMT) and median, respectively. The cut-off values were represented by dotted lines.

### Increase binding antibodies level after boost

Waning immunity against SARS-CoV-2 in heterologous CoronaVac/AZD1222 primed individuals was determined. The anti-RBD IgG at one month (n = 35) as reported in a previous study [23] was individually compared to 4 to 5 months after the second dose (baseline in this cohort) as shown in Supplementary Fiigure 1. As expected, a significant drop (5.9-fold) in anti-RBD IgG within 4 to 5 months (109.6 BAU/mL) occurred compared to one-month post-second dose (652.1 BAU/mL; *p* < 0.001). This result indicates a decline in immunity over time in vaccinated individuals who received heterologous CoronaVac/AZD1222. Following booster vaccination, the anti-RBD IgG significantly increased and peaked at day 14 for all vaccines (*p* < 0.001) as shown in Figure 1B. Comparing pre- and post-boost, mRNA-1273-boosted individuals achieved an anti-RBD IgG with a 23-fold increase (126.9 versus 2921 BAU/mL) and showed a higher level than the other vaccine groups, while BNT162b2 and AZD1222 groups were induced by 15.8-fold (152.1 versus 2404 BAU/mL) and 2.5-fold (142 versus 360.8 BAU/mL) higher than baseline. Furthermore, anti-RBD IgG levels correlated negatively with age in boosted mRNA-1273 individuals although these differences were not significant. On the contrary, the age-related trend was not found in AZD1222- or BNT162b2-boosted individuals (Figure 1C). Most boosted individuals were seronegative for anti-N IgG, indicating no SARS-CoV-2 exposure during the study period (Figure 1D). Although one participant had an anti-N IgG level above the cut-off, the anti-RBD IgG level was comparable with other participants at baseline, suggesting the anti-N IgG may be induced by CoronaVac prime vaccination.

### Neutralizing activity to delta and omicron measured using sVNT

Neutralizing activity to the delta and omicron variants was measured in sera at baseline and 28 days after post-boost using the surrogate virus neutralization test. All boosted individuals could restore neutralizing activity to delta by more than 90% (Figure 2A). Although baseline neutralizing activity to omicron declined at 4 to 5 months after the second dose, a 20% (6/30), 83% (25/30), and 90% (27/30) of individuals boosted with AZD1222, BNT162b2, and mRNA-1273, respectively, were detected to possess omicron variant neutralizing potential (Figure 2B). By comparison, it was noted that individuals boosted with mRNA vaccines demonstrated a higher level of neutralizing activity than those boosted with AZD1222. The median of neutralizing activity to omicron was 10.1% for AZD1222, 55.9% for BNT162b2, and 78.2% for mRNA-1273 after booster vaccination. Although neutralizing activity against omicron was significantly lower than that against the delta variant, most individuals have detected the neutralizing activity against omicron after receiving booster mRNA vaccines (Figure 2C).

**Figure 2:**
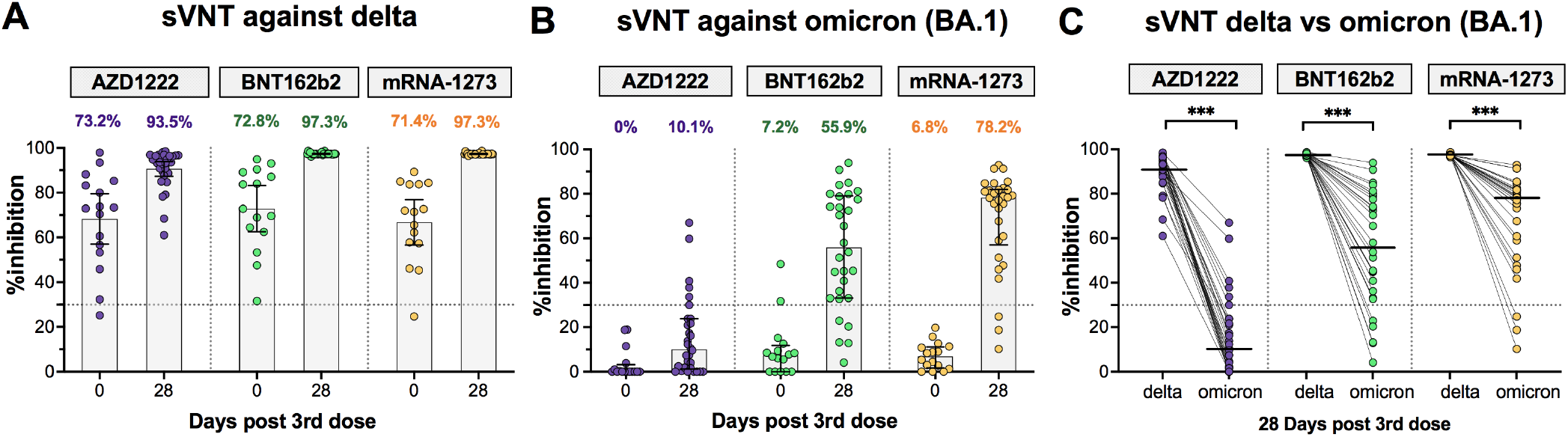
Neutralizing activities compared between pre-and post-boost measured using surrogate virus neutralization test. A subset of samples from boosted individuals with AZD1222 (purple), BNT162b2 (green) and mRNA-1273 (yellow) that was randomly selected to test the surrogate virus neutralization test (sVNT) that included sera collected at baseline (n=15/group) and sera collected at 28 days post-boost (n=30/group). **Panel A** shows neutralizing activity against delta and **Panel B** shows neutralizing activity against omicron (BA.1). Numbers above the bar graph indicate the percentage of inhibition between human angiotensin converting enzyme 2 and receptor binding domain (ACE-2 and RBD, respectively) proteins. **Panel C** shows a comparison of the neutralizing activity between delta and omicron variants after booster vaccination. Median values with interquartile ranges (IQRs) are shown as horizontal bars. Dotted lines indicate cut-off values (30%). The comparison was perform using Wilcoxon signed-rank test (two-tailed). ***, p < 0.001.

### Comparison of neutralizing antibody titers to omicron BA.1 and BA.2

The functional NAb titers against omicron BA.1 and BA.2 were quantified using a live virus neutralization test (FRNT50). At baseline, 80% (24/30) and 43% (13/30) of vaccinated individuals with heterologous CoronaVac/AZD1222 had NAbs to omicron BA.1 and BA.2, respectively, which dropped below detectable levels (Figure 3). After 28 days post-boost, Nab titers against omicron BA.1 were 40.34, 171.0, and 271.6 after AZD1222, BNT162b2, and mRNA-1273 boosters, respectively, reflecting a 3.16-, 9.91- and 24.78-fold increase compared to baseline, respectively (Figure 3A). The NAb titers against omicron BA.2 in AZD1222, BNT162b2, and mRNA-1273 groups were 59.27, 130.7, and 235.3, respectively, which reached a 2.43-, 4.63- and 19.67-fold induction relative to baseline, respectively (Figure 3B). This finding indicates that the omicron variant is more susceptible to neutralization by sera from individuals boosted with mRNA vaccines (BNT162b2 and mRNA-1273) than those boosted with AZD1222. Overall, the NAb titers to BA.1 and BA.2 were comparable. By comparison, the NAb titer to BA.2 was 1.47-fold higher in the AZD1222 group and 1.31- and 1.15-fold lower in the BNT162b2 and mRNA-1273 groups, respectively, compared to BA.1 (Figure 3C). Moreover, it was shown that anti-RBD IgG and sVNT to omicron correlated well with the NAb titers against omicron BA.1. and BA.2 as measured using the live virus neutralization test (FRNT50) as shown in Supplementary Fiigure 2.

**Figure 3.**
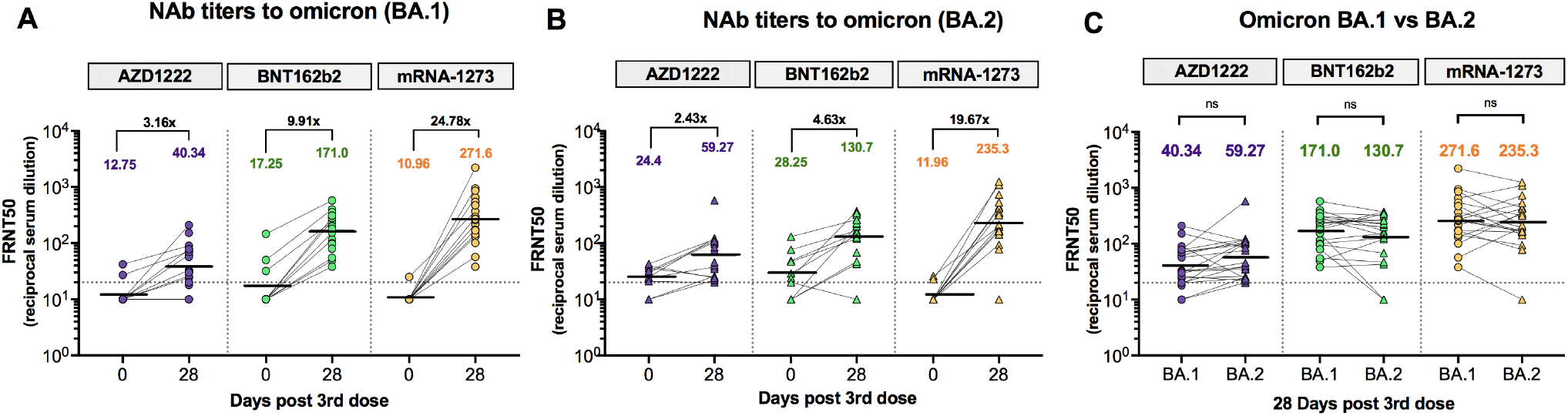
Neutralizing antibody titers against omicron BA.1 and BA.2 measured using focus reduction neutralization test (FRNT50). A subset of samples from boosted individuals with AZD1222 (purple), BNT162b2 (green) and mRNA-1273 (yellow) that was randomly selected to test the FRNT50 included sera collected at baseline (n = 10/group) and sera collected at 28 days post-boost (n = 30/group). NAb titers against omicron BA.1 **(Panel A)** and omicron BA.2 **(Panel B)** were compared between baseline (day 0) and 28 days post-booster vaccination with different vaccines. Fold-increases for each comparison are denoted. NAb titers against BA.1 and BA.2 after booster vaccination were compared (**Panel C**). Statistical analysis was done using Wilcoxon signed rank test (two-tailed). The horizontal dotted line indicates the limit of detectable value of FRNT50. Values below the limit of detection (< 20) were set at a titer of 10 before statistical analysis. ns indicates no significant difference.

### Total interferon-gamma response

Besides the neutralizing antibodies, the T cell response were assessed by measuring total IFN-γ responses in whole blood from AZD1222-, BNT162b2-, and mRNA-1273-boosted individuals after S1 (RBD) peptides for CD4+ stimulation or Ag1 (Figure 4A) and S1+S2 peptides for CD4+ and CD8+ stimulation or Ag2 (Figure 4B). At baseline, 47% (43/90) and 57.7% (52/90) of participants could elicit IFN-γ responses after Ag1 and Ag2 stimulation after 4 to 5 months post-second dose. Following booster vaccination, IFN-γ levels significantly increased at 14 days after receiving BNT162b2 and mRNA-1273 as booster doses. Notably, 90%–97% of individuals boosted with BNT162b2 and mRNA-1273 could induce IFN-γ responses after stimulation with the spike protein of the ancestral strain. On the contrary, no difference in IFN-γ response in those boosted with AZD1222 compared to baseline was found. This result indicates that individuals boosted with BNT162b2 and mRNA-1273 vaccines could induce a T-cell response, which elicits a higher level of IFN-γ response, but this process was not observed in those boosted with the AZD1222 vaccine.

**Figure 4:**
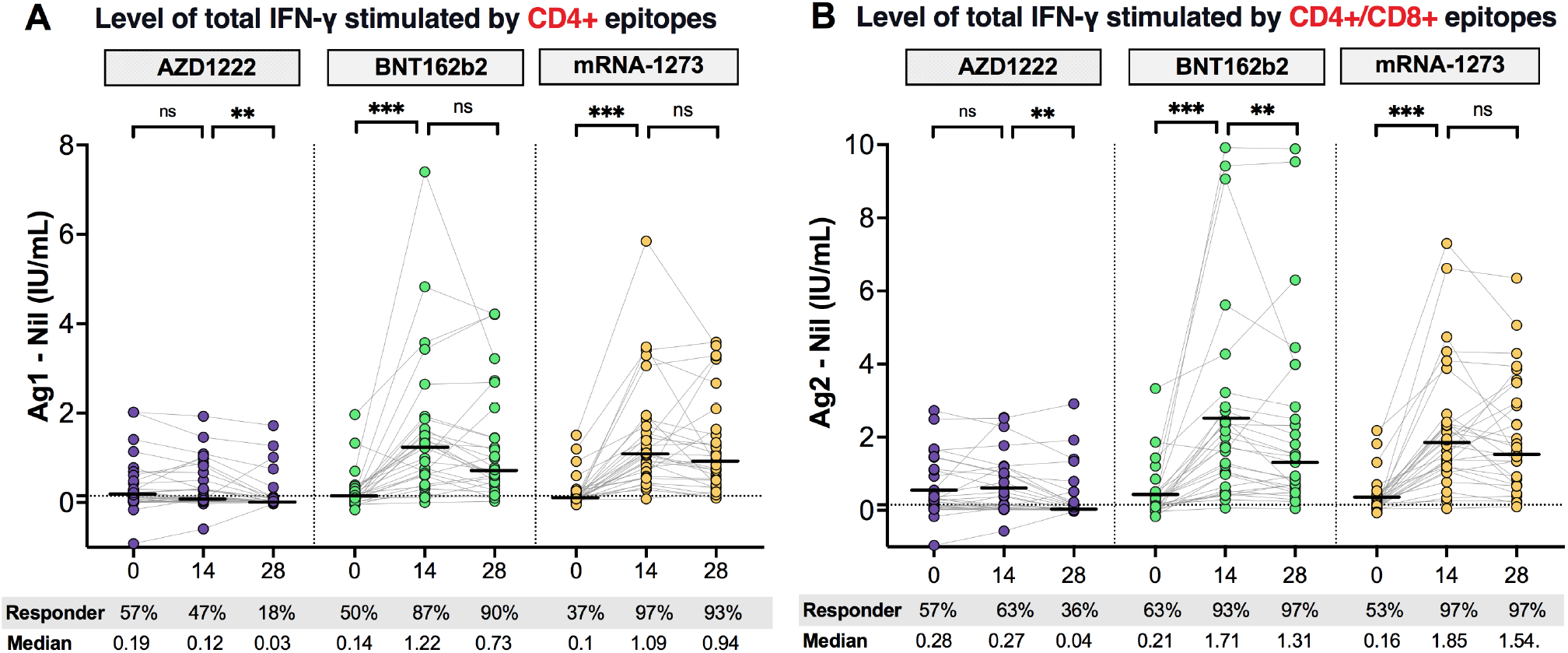
Comparison of total IFN-γ -releasing T-cell responses to SARS-CoV-2 antigens. Serum samples from vaccinated individuals receiving heterologous CoronaVac/AZD1222 followed by a third booster with AZD1222 (*n* = 30, purple), BNT162b2 (*n* = 30, green), or mRNA-1273 (*n* = 30, yellow) were stimulated by (A) Ag1, which is a CD4+ epitope derived from RBD, minus negative control (Nil), and (B) Ag2, which is CD4+ and CD8+ epitopes derived from S1 and S2 subunits, minus negative control (Nil). Levels of IFN-γ above cutoff values (0.15 IU/mL and ≥ 25% of Nil) indicate a reactive response. Horizontal bars indicate the median. The cut-off values were represented by horizontal dotted line. Statistical analysis was done using Wilcoxon signed rank test (two-tailed). ns indicates no significant difference; **, p < 0.01; ***, p < 0.001.

### Reactogenicity after booster vaccination

Local and systemic reactogenicity were self-reported within seven days after booster vaccination. A high frequency of adverse events (AEs) was observed within 2 to 3 days following the booster dose and were predominantly mild to moderate (Supplementary Fiigure 3). Boosted individuals with BNT162b2 and mRNA-1273 vaccines reported greater local and systemic reactogenicity than those receiving the AZD1222 vaccine. Overall, the most common AEs observed in boosted individuals were injection site pain, redness, and swelling indicating local AEs, while myalgia headache and chills were frequently reported as systemic AEs (Figure 5). However, no serious AEs were reported.

**Figure 5:**
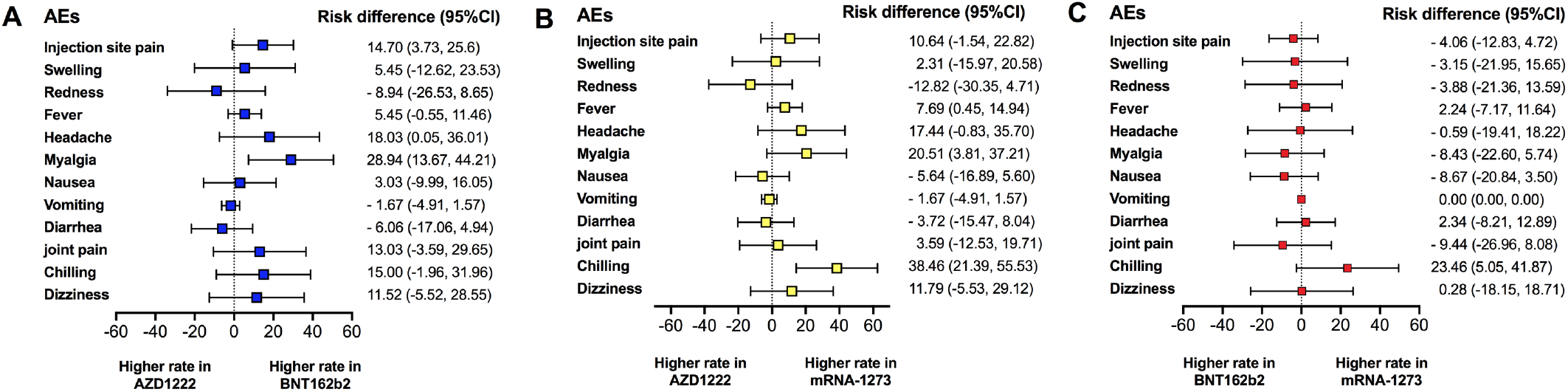
Forest plot showed the risk difference with 95% confidence intervals od adverse events (AEs) after the booster dose. The proportion of participants with any grade of solicited Aes after the third booster were compared between **(Panel A)** AZD1222 versus BNT162b2 vaccine **(Panel B)** AZD1222 versus mRNA-1273, and **(Panel C)** BNT162b2 versus mRNA-1273.

## DISCUSSION

This study demonstrated the NAb response against omicron BA.1 and BA.2 subvariants and T-cell responses after boosting with AZD1222, BNT162b2, and mRNA-1273 in vaccinated individuals who had received heterologous CoronaVac/AZD1222 prime. It was found that a booster vaccination could restore the binding antibody response and induce NAb titers against omicron BA.1 and BA.2. Of note, our findings indicate that individuals boosted with mRNA-1273 and BNT162b2 vaccines could induce humoral and T-cell responses higher than those boosted with AZD1222. Although mRNA vaccines showed a higher frequency of AEs than AZD1222 vaccine, the reactogenicity was in acceptable ranges, indicating a good safety profiles after booster vaccination.

It was found that individuals who received the heterologous CoronaVac/AZD1222 vaccination exhibited a 5.9-fold reduction in binding antibody and less detectable NAbs to omicron variants after 4 to 5 months post-second dose, indicating waning of vaccine-induced immunity [10]. It has been well established that booster vaccinations could overcome the waning immunity [14,21,22,28]. Rapid elevation in binding antibodies after boosting with mRNA and adenoviral-vectored vaccines were found. Similar results have been reported after receiving either a viral-vectored or an mRNA vaccine as a third dose, indicating that it was sufficient to recall the memory B-cells [28]. Typically, vaccine-induce immunity negatively correlates with age and drops significantly around the ages of 80 years [29]. However, those age-related trends were not found in this study, which might because most participants were less than 60 years old.

Neutralizing antibody titers is a highly predict the immune protection against SARS-CoV-2 infection. Higher levels of NAbs correlated with a reduced risk of infection and severe disease [11]. Our study found a substantial loss of NAb titers against omicron variants at pre-booster vaccination. This effect might be related to harboring of mutations concentrated around the RBD [3]. However, the NAb titers against omicron were retained upon booster vaccination even though the current vaccines composition was based on the ancestral strain. Consistent results were observed after receiving mRNA vaccine following either homologous inactivated, adenoviral-vectored, or mRNA-based vaccines prime [14,21,22,30]. Although the low number of omicron-neutralizing memory B-cells produced after two-doses of mRNA vaccination has previously been reported, their coverages were improved through affinity maturation over time and might be sufficient to expand the coverage of neutralization against SARS-CoV-2 variants after receiving a booster dose [31]. Furthermore, a reduced neutralizing activity against omicron compared to delta variant was found. Similar results were found concerning neutralizing activity to omicron, which was reduced by 6-to 23-fold lower than delta after booster vaccination [12].

With the surge of omicron BA.1 and BA.2 subvariants, a substantial loss of NAbs to omicron BA.1 and BA.2 subvariants was observed in vaccinated individuals with two doses of mRNA vaccine and patients who previously were infected with wild-type SARS-CoV-2 [32]. Furthermore, a study of the neutralizing antibody of omicron subvariants indicated that a 23-fold reduction for BA.1 and a 27-fold reduction for BA.2 was observed compared to wild -type in vaccinated individuals who received the two doses of BNT162b2 vaccine [30]. Our findings indicate that booster vaccination with mRNA and adenoviral-vectored vaccines could increase the neutralizing antibody titers and coverage against omicron BA.1 and BA.2. Consistent results showed an improvement in neutralization sensitivity against BA.1 and BA.2 after individuals were boosted with the mRNA vaccine [30,33]. Although both subvariants shared several common amino acid changes, the unique mutations found in each subvariant might affect the differences in neutralization potency [32]. However, it was found that the NAb titer of BA.1 and BA.2 were comparable and trended higher in the adenoviral-vectored booster and lower in mRNA vaccines. A recent study showed a 1.4-fold lower neutralizing antibody titer to BA.2 compared to BA.1 [30]. This result indicates that booster vaccination is a useful strategy for controlling the omicron BA.1 and BA.2 pandemic.

A study of T-cell responses targeting the omicron spike protein suggested that no differences in T-cell profiles and cytokine production between omicron and wild type were found [17]. This finding indicates cross-recognition of T-cells was minimally affected by mutations in the omicron spike protein [18,34]. A current study showed that individuals boosted with the mRNA vaccine could induce T-cell activity in whole blood. Cross-recognition of different SARS-CoV-2 variants by T-cells was maintained after being boosted with mRNA vaccine has been reported [17,35]. On the contrary, our result showed that individuals boosted with AZD1222 following heterologous CoronaVac/AZD1222 appeared to abolish T-cell responses.

Study limitations include the detection limits of the surrogate virus neutralization assay. Most boosted individuals elicited an elevation in antibody level that was higher than the upper limit of detection; thus, this method may not have provided the actual neutralizing antibody level in case of a robust immune response. Furthermore, the effect of omicron peptide stimulation on T-cell response was not examined. However, van Kessel et al. showed that no difference in cytokine production was observed upon stimulation with S-peptide pools derived from the ancestral strain and omicron variants [17]. Further studies are required to define long-term immunity and durability of immune responses against SARS-CoV-2 variants, particularly omicron subvariants, after administration of booster vaccinations.

In conclusion, these findings indicate that booster vaccination could retain the level of anti-RBD IgG and improved the neutralizing activity against delta and omicron variants. Of note, a booster dose could induce neutralizing antibody titers to omicron BA.1 comparable to BA.2. Furthermore, individuals boosted with mRNA vaccines could induce IFN-γ responses higher than those boosted with AZD1222. Hence, giving mRNA vaccines as the booster dose could improve humoral and T-cell responses and induce neutralization coverage against omicron subvariants.

## Data Availability

All data produced in the present work are contained in the manuscript

## Acknowledgements

We would like to thank all the staffs of the Center of Excellence in Clinical Virology and all the participants for helping and supporting in this project. We also appreciate the Ministry of Public Health, Chulabhorn Royal Academy and Zullig pharma for providing the vaccines for this study.

## Corresponding author contact information

1. Prof. Dr. Yong Poovorawan, Center of Excellence in Clinical Virology, Department of Pediatrics, Faculty of Medicine, Chulalongkorn University. Bangkok, 10330 Thailand.

