## Supplementary Information for "Effects of boosted mRNA and adenoviral-vectored vaccines on immune responses to omicron BA.1 and BA.2 following the heterologous CoronaVac/AZD1222 vaccination"

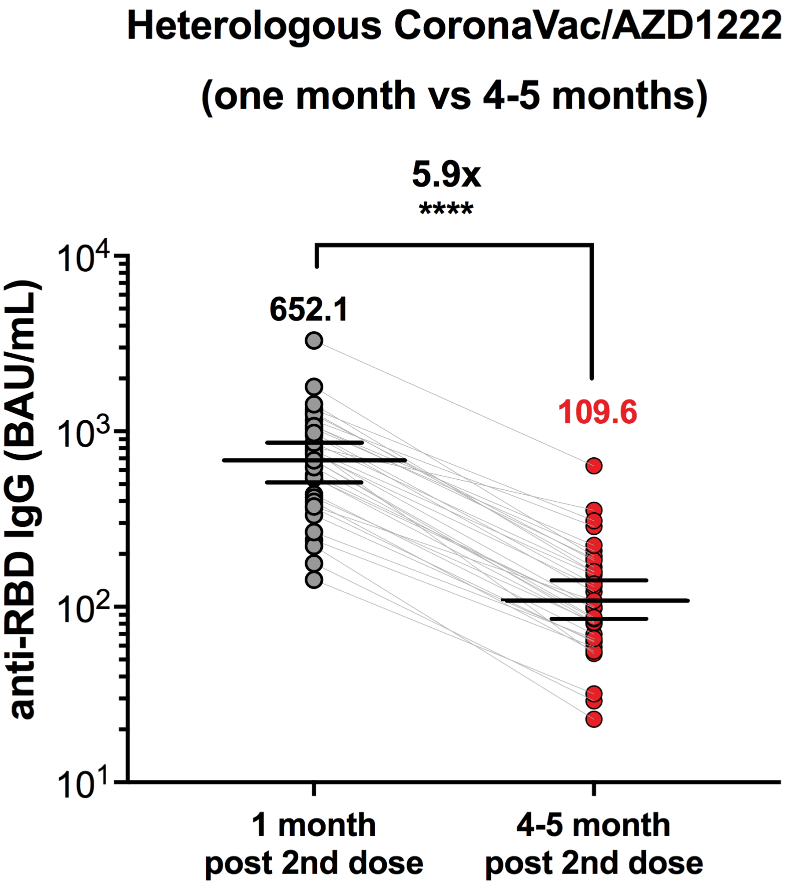


**Supplementary Figure 1.** Vaccine-induced immunity wane in individuals who received heterologous CoronaVac/AZD1222 vaccination. Pair serum samples (n=35) at one-month post-vaccination obtained from our previous report [23] were compared to 4-5 months post-vaccination from current study. Numbers above the column indicate the geometric mean titers (GMT). Error bars indicate the GMT with 95% CIs and fold-reduction is denoted. The comparison was performed using a t-test based on log-transformed data. ***p < 0.001.

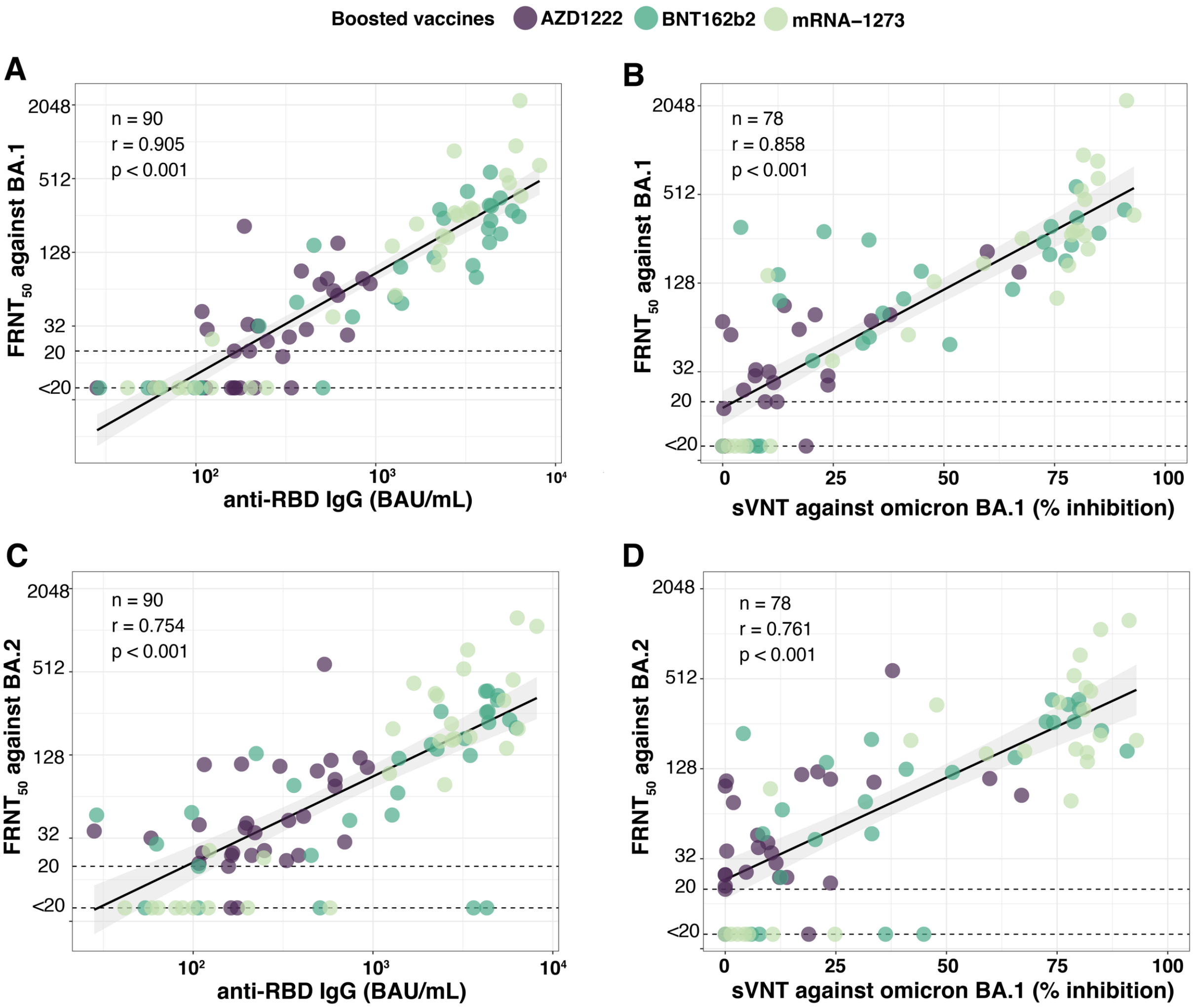

Boosted vaccines
AZD1222
BNT162b2
mRNA−1273
A
n = 90
r = 0.905
p < 0.001
FRNT50 against BA.1
anti-RBD IgG (BAU/mL)
B
n = 78
r = 0.858
p < 0.001
FRNT50 against BA.1
sVNT against omicron BA.1 (% inhibition)
C
n = 90
r = 0.754
p < 0.001
FRNT50 against BA.2
anti-RBD IgG (BAU/mL)
D
n = 78
r = 0.761
p < 0.001
FRNT50 against BA.2
sVNT against omicron BA.1 (% inhibition)

**Supplementary Figure 2**. Correlation between neutralizing antibody titers against omicron subvariants tested using a live virus neutralization test (FRNT50) and serological testing. Correlation between FRNT50 against BA.1 and anti-RBD IgG (RBD from the ancestral strain) (Panel A), FRNT50 against BA.1 and sVNT against BA.1 (Panel B), FRNT50 against BA.2 and anti-RBD IgG (Panel C), FRNT50 against BA.2 and sVNT against BA.1 (Panel D) were analyzed. For neutralizing antibody titers (FRNT50 titers), the value <20 was defined as undetectable and set as a FRNT50 of 10 (dotted lines). For anti-RBD IgG, the value ≥ 7.1 BAU/mL was defined as a positive response. The cut-off for sVNT was 30 % inhibition. Spearman rank correlation test was calculated based on log-transform data. BAU = binding arbitrary units, FRNT50 = foci reduction neutralization titer – 50%.

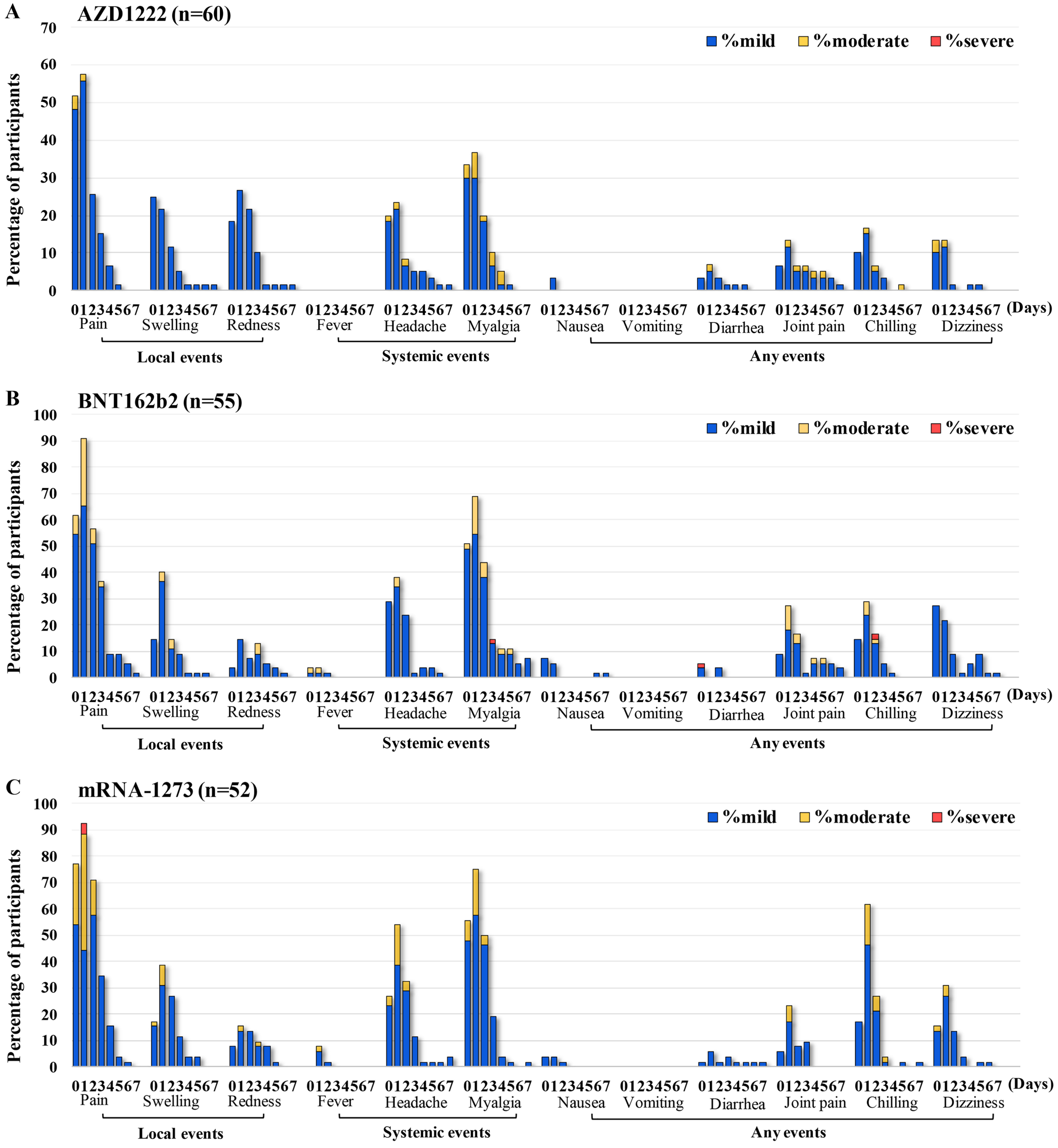


**Supplementary Figure 3. Solicited local and systemic adverse events within seven days after third dose vaccination in individuals who previously received heterologous CoronaVac/AZD1222 vaccination.** Bar graphs present adverse events following the third dose vaccination in 167 participants who received AZD1222 (*n* = 60) **(Panel A),** BNT162b2 (*n* = 55)

**(Panel B)** and mRNA-1273 (*n*=52) **(Panel C)**. Day 1 is the day of the third dose of vaccination. Fever was classified as mild (38 °C to <38.5 °C), moderate (38.5 °C to <39 °C), and severe (≥39 °C). The swelling was graded as mild (<5 cm), moderate (5 cm to <10 cm), and severe (≥10 cm). Symptoms were graded as follows: mild, no limitation on normal activity; moderate, some limitation of daily activity; and severe, unable to perform or daily activity prevented.
